# SARS-CoV-2 Viral Load Predicts COVID-19 Mortality

**DOI:** 10.1101/2020.06.11.20128934

**Authors:** Elisabet Pujadas, Fayzan Chaudhry, Russell McBride, Felix Richter, Shan Zhao, Ania Wajnberg, Girish Nadkarni, Benjamin Glicksberg, Jane Houldsworth, Carlos Cordon-Cardo

**Author notes:** Corresponding author: Carlos Cordon-Cardo, MD, PhD. These authors contributed equally (Elisabet Pujadas, Fayzan Chaudhry).

## Abstract

The need for reliable and widely available SARS-CoV-2 testing is well recognized, but it will be equally necessary to develop quantitative methods that determine viral load in order to guide patient triage and medical decision making. We are the first to report that SARS-CoV-2 viral load at the time of presentation is an independent predictor of COVID-19 mortality in a large patient cohort (n=1,145). Viral loads should be used to identify higher-risk patients that may require more aggressive care and should be included as a key biomarker in the development of predictive algorithms.

## Introduction

SARS-CoV-2 detection platforms currently report qualitative results. However, RT-PCR-based technology allows for quantification of viral loads, which have been associated with transmission risk and disease severity in other viral illnesses^1^. Similarly, viral load in COVID-19 may correlate with infectivity, disease phenotype, morbidity and mortality. To date, few studies have reported on SARS-CoV-2 viral loads, and no studies have assessed viral load and mortality in large patient cohorts.^2,3,4,5^ This letter is the first to report SARS-CoV-2 viral load at the time of diagnosis as an independent predictor of COVID-19 mortality in a large hospitalized cohort (n*=1,145*) within the Mount Sinai Health System in New York City.

## Methods

We prospectively evaluated nasopharyngeal swab samples for SARS-CoV-2 by the Roche cobas® 6800. Positive samples were also evaluated by a laboratory-developed quantitative RT-PCR test^6^. A six-fold standard dilution curve was run in triplicate (limit of detection 1×10^3^ viral RNA genome equivalents/mL). Viral loads (N2) were log transformed and a constant was added.

## Results

Viral loads for 1,145 hospitalized SARS-CoV-2 positive patients were measured on samples collected between 3/13/2020 and 5/4/2020 that tested positive on both the Roche cobas® 6800 and the laboratory-developed test at the time of diagnosis. The cohort had an average age of 64.6 years, was 56.9% male, and had a self-reported race breakdown of African-American (31.2%), White (29.3%), Asian (3.7%), Other (32.8%) and Unknown (3.1%). The overall mean log10 viral load for the group was 5.56 viral copies/mL, and the median log10 viral load was 6.16 viral copies/mL. By the end of the study period, 807 were alive (70.5%; mean log10 viral load 5.19 +/- 2.99 viral copies/mL) and 338 had died (29.5%; mean log10 viral load was 6.44 +/- 2.66 viral copies/mL).

A Cox proportional hazards model was performed to evaluate the association between viral load and mortality, adjusting for multiple baseline clinical and demographic characteristics including age, sex, race, asthma, atrial fibrillation, coronary artery disease, chronic kidney disease, chronic obstructive pulmonary disease, heart failure, hypertension, and stroke that yielded a statistically significant independent association between viral load and mortality (HR 1.069, CI 1.026-1.11; p=0.0014) (**Figure 1A**). Furthermore, a univariate survival analysis revealed a statistically significant survival probability between those with a high (defined as greater than the log10 viral load mean of 5.557) and low viral load (p = 0.0003; **Figure 1B**), with a mean follow up time of 12.8 days, and a maximum follow up of 66 days.

**Figure 1.**
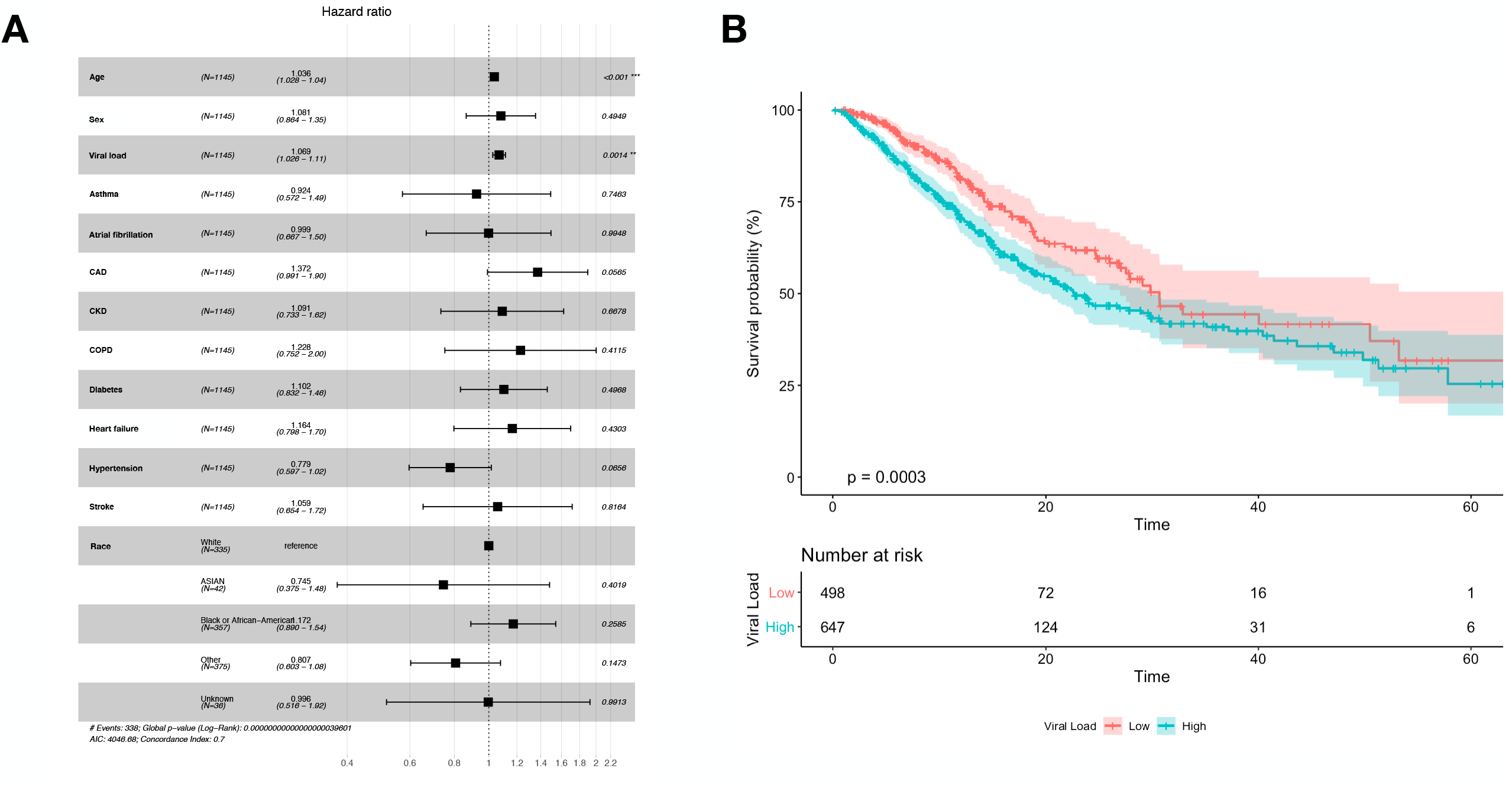
Viral load is an independent predictor of SARS-CoV-2 related mortality. **(Part A)** Cox proportional hazards model evaluating the association between viral load and mortality, adjusting for multiple baseline clinical and demographic characteristics. **(Part B)** Univariate survival analysis showing a statistically significant survival probability between those with a high and low viral load.

## Discussion

Deployment and scaling of testing have been critical in the management of COVID-19, yet the challenge of understanding who is at risk for worse outcomes early in their illness remains. Here, we demonstrate a relationship between high viral load and increased mortality in a large hospitalized cohort. High viral load could signify greater disease severity or a more compromised host, with both scenarios requiring closer clinical monitoring and a high index of suspicion for poor outcome and death.

Transforming front-line qualitative testing into a quantitative viral load will assist clinicians in risk-stratifying patients who should be admitted and closely monitored versus those who may safely convalesce at home. Viral loads may also affect isolation measures based on infectivity. It will also be of great interest to address SARS-CoV-2 viral load dynamics in individual patients and the quantitative relationship with neutralizing antibodies, cytokines and other covariates.

## Data Availability

Data available upon request provided all appropriate patient and data protection criteria are met.

